# Cumulative Multidomain Vulnerability and Early Mortality After Heart Transplantation: Development and Temporal Evaluation of the RIGHT-HTx Framework

**DOI:** 10.64898/2026.11.03.26367231

**Authors:** Komarakshi Balakrishnan, Rao K.G. Suresh, Sahayaraj R. Anto, Murali Krishna Tanguturu, R.K.R. Noveen Davidson, Ajay Aravind, Sudheer Babu Vanga, A. Jagadish, Nithya Nallamuthu, Nehal C. Singh, Vinod Manikala, A. Arun Kumar, K.G. Krithika, Ramyashri Chandrasekaran, R. Rajesh Kumar, Ravikumar Ratnagiri

## Abstract

**BACKGROUND:** Risk models from high-income transplant programmes may not generalise to resource-constrained settings with different recipient acuity, donor characteristics and access to mechanical circulatory support. We developed RIGHT-HTx, a parsimonious framework integrating clinical and physiological vulnerabilities, and evaluated its prognostic performance in a subsequent same-centre cohort.

**METHODS:** This retrospective single-centre study included 436 consecutive adult heart transplant recipients (2012– 2026). Of 357 development recipients, 341 (95.5%) were score-evaluable. RIGHT-HTx incorporated three vulnerability domains: clinical acuity (INTERMACS 1–2), graded renal dysfunction (eGFR 30– 59 or <30 mL/min/1.73 m2) and haemodynamic–donor mismatch (TPG >12 mmHg combined with donor-to-recipient weight ratio <1.0). A seven-point Cox-derived score underwent internal validation using 1,000 bootstrap resamples and was applied unchanged to 79 subsequent recipients. Low-, intermediate- and high-risk categories represented zero, one and ≥2 affected domains, respectively.

**RESULTS:** Among development recipients, 75/341 (22.0%) died within one year. Mortality increased across low-, intermediate- and high-risk categories (8.1%, 20.3% and 55.6%; log-rank P<0.001). Elevated TPG was not significantly associated with mortality individually, whereas its combination with donor undersizing was independently associated with mortality (adjusted HR 2.87, 95% CI 1.53–5.40; P=0.001). The full model improved discrimination beyond renal dysfunction alone (optimism-corrected ΔC-index 0.067), and the simplified score achieved an optimism-corrected C-index of 0.732.

The temporal cohort more often had ≥1 affected domain (75.9% versus 60.4%; P=0.010). Discrimination was comparable (C-index 0.738, 95% CI 0.643–0.833), although observed Kaplan– Meier one-year mortality exceeded original predictions (35.4% versus 25.4%; calibration slope 0.929). In pooled analyses (n=420), mortality increased from 8.4% (13/154) with no affected domain to 57.3% (47/82) with two or more.

**CONCLUSIONS:** RIGHT-HTx identifies cumulative multidomain vulnerability associated with early mortality after heart transplantation. Temporal discrimination was comparable, but absolute-risk calibration differed. The framework may support risk recognition and mitigation, not transplant eligibility or allocation; independent multicentre validation is required before clinical implementation.

## INTRODUCTION

Heart transplantation remains the definitive treatment for selected patients with end-stage heart failure.^1^ Most transplants and published risk models originate in North America and Europe.^2,3,4^ Their transportability to programmes with different recipients, donor pools, and access to mechanical circulatory support (MCS) is uncertain. External validation and calibration assessment are frequently incomplete,^5^ and model performance may vary across populations^6^ and between scores evaluated in the same transplant cohort.^7^

These concerns are especially pertinent to resource-constrained programmes, where recipients may rely on personal funds,^8^ durable MCS access is limited,^9,10^ and deceased donors are geographically concentrated. Critically ill patients must therefore endure long-distance travel and prolonged waiting periods without durable mechanical support.^11^ Heart transplantation also depends on a scarce donor resource beyond the treating team’s control. Socioeconomic and geographical factors influence heart-failure and transplant outcomes,^12,13,14^ alongside regional differences in transplant survival.^2^ Nevertheless, the underlying physiological determinants of early mortality are likely to be relevant across healthcare systems, even when the resources available to address them differ.

We sought to identify clinically relevant predictors of one-year mortality following heart transplantation in a resource-constrained setting and to develop a parsimonious, clinically interpretable risk-stratification framework. Candidate predictors were examined using clinical and physiological considerations alongside survival analyses, from which the RIGHT-HTx score was derived. Its performance was subsequently evaluated in a non-overlapping, same-centre temporal cohort. The score is intended to support risk recognition and potential mitigation, not determine transplant eligibility or organ allocation.

## MATERIALS AND METHODS

### Study Design and Population

This retrospective, single-centre study included 357 consecutive adult heart transplant recipients transplanted between October 2012 and September 2024 and 79 subsequent recipients transplanted between October 2024 and June 2026. The latter constituted a non-overlapping, same-centre temporal cohort. Paediatric and combined-organ transplant recipients were excluded.

Of 357 development recipients, 341 (95.5%) were score-evaluable; five lacked estimated glomerular filtration rate (eGFR), and 11 had indeterminate haemodynamic–donor classification. All 79 temporal recipients were evaluable, yielding 420 recipients for pooled analyses. Complete-case analysis was used for model development without multiple imputation. Mortality among score-evaluable and non-evaluable recipients was compared to assess potential selection bias.

The primary endpoint was all-cause mortality within 12 months of transplantation, analysed using time-to-event methods with administrative censoring at one year. Survival status was updated through 25 October 2026. No temporal recipient was lost to follow-up; 17 had less than 12 months of observation and were censored at their last confirmed contact. Five score-evaluable development recipients were also censored before 12 months at their last documented follow-up.

Clinical, biochemical, haemodynamic, donor and transplant data were obtained from prospectively maintained institutional electronic records. The Institutional Review Board approved the study and waived individual informed consent. Reporting followed TRIPOD+AI guidance.^15^

### Variables and Definitions

eGFR was calculated using the 2021 race-free CKD-EPI creatinine equation.^16^ MELD-XI was calculated as 5.11 × ln(bilirubin) + 11.76 × ln(creatinine) + 9.44, with values below 1 mg/dL set to 1.17

Clinical acuity was classified by INTERMACS profile at transplantation rather than listing, reflecting deterioration while awaiting transplantation where applicable.^18^ Recipients requiring extracorporeal membrane oxygenation or temporary mechanical support for critical cardiogenic shock were classified as profile 1. Durable-LVAD recipients were reclassified as profile 1 only following documented clinical deterioration. Profiles were dichotomised as 1–2 versus 3–5.

Right-heart catheterisation was performed within three months before transplantation, but the exact interval between right-heart catheterisation and transplantation was not recorded. Transpulmonary gradient (TPG) was calculated as mean pulmonary arterial pressure minus pulmonary capillary wedge pressure; pulmonary vascular resistance (PVR) as TPG divided by cardiac output. Recorded haemodynamic indices were retained when constituent measurements were unavailable.

Donor undersizing was defined as donor-to-recipient body-weight ratio <1.0. Haemodynamic–donor vulnerability required both TPG >12 mmHg and donor undersizing. The composite was classified as absent in non-undersized recipients regardless of TPG availability; TPG was required for classification when donors were undersized.

### RIGHT-HTx Development and Scoring

Candidate predictors were examined using univariable Cox regression. Selection prioritised clinical relevance, physiological plausibility, nonredundancy and parsimony, informed by observed associations. The RIGHT-HTx framework incorporated three vulnerability domains: INTERMACS-defined clinical acuity, renal dysfunction and combined haemodynamic–donor mismatch.

The final multivariable Cox model contained four coefficients: INTERMACS 1–2, eGFR 30–59 and <30 mL/min/1.73 m2 (reference ≥60), and TPG >12 mmHg with donor undersizing. RIGHT-HTx represents Renal reserve, INTERMACS-defined acuity, Graft–recipient size compatibility and Haemodynamic load in heart Transplantation; the latter two were combined into one model term.

Regression coefficients were converted to integer points by dividing by half the smallest nonreference coefficient (0.8803/2) and rounding, following established methods for presenting multivariable models as integer point scores.19 Points were allocated as follows: INTERMACS 1–2, 2; eGFR 30–59, 2; eGFR <30, 3; and haemodynamic–donor mismatch, 2. The maximum score was seven.

Risk categories reflected the number of affected domains: low (none; score 0), intermediate (one; scores 2–3) and high (two or more; scores 4–7). Category boundaries followed the domain structure rather than outcome-optimised cutoffs. Donor age and ischaemic time were evaluated but not retained.

### Incremental Prognostic Value and Internal Validation

Incremental prognostic information was assessed by comparing renal-only, renal-plus-INTERMACS and full three-domain Cox models using nested likelihood-ratio tests and Harrell’s C-index. Reverse-order comparisons examined the contribution of each additional domain.

Internal validation used 1,000 bootstrap resamples. Full-model coefficients were re-estimated within resamples, while the simplified seven-point score was evaluated with fixed integer weights and a re-estimated score coefficient. Optimism-corrected discrimination and calibration slopes were calculated. A separate paired bootstrap comparison estimated the optimism-corrected improvement in discrimination over renal dysfunction alone.

Validation was conditional on the selected predictors, thresholds, weights and risk boundaries; the complete development process was not repeated. Absolute 12-month risks were estimated using the continuous seven-point Cox model, with calibration assessed against Kaplan–Meier mortality.

### Haemodynamic–Donor Interaction and Sensitivity Analyses

Recipients were classified into four groups according to TPG ≤12 or >12 mmHg and donor undersizing. An analogous analysis used PVR ≤3 or >3 Wood units. Cox models examined these groups and multiplicative interactions, adjusting for INTERMACS status and eGFR categories. Interaction terms were evaluated using nested likelihood-ratio tests.

The TPG and PVR analyses included 312 and 313 recipients, respectively; adjusted interaction models included 309. An additional complete-case sensitivity analysis compared the incremental prognostic contributions of MELD-XI and eGFR categories using nested Cox models fitted to a common sample. These analyses were exploratory, did not constitute independent validation and were not adjusted for multiplicity.

### Temporal Evaluation and Cumulative Vulnerability

The unchanged seven-point score was applied to 79 subsequent recipients. Discrimination was assessed using Harrell’s C-index, and calibration by comparing frozen development-model predictions with observed Kaplan–Meier 12-month mortality. The calibration slope was estimated using the frozen linear predictor.

Exploratory recalibration retained the development score coefficient while updating the baseline cumulative hazard using temporal outcomes; these predictions were not considered independent validation.

For descriptive pooled analyses, 420 score-evaluable recipients were classified by the number of affected domains (0 to 3). Renal dysfunction counted as one domain regardless of eGFR severity. Crude mortality proportions and Wilson confidence intervals were calculated. These pooled analyses were not independent of model development.

### Long-Term and Conditional Survival

Programme-level survival was assessed in all 436 recipients using Kaplan–Meier methods. Conditional survival was evaluated among development-cohort recipients surviving and remaining under observation beyond one year, retaining their original RIGHT-HTx classifications. Between-group differences were assessed using log-rank tests and Cox regression.

### Statistical Analysis

Continuous variables were summarised as median (interquartile range) and categorical variables as number (percentage). Cox models used the Efron method for ties, with hazard ratios and 95% confidence intervals reported. Proportional-hazards assumptions were assessed using Schoenfeld residuals. Two-sided P<0.05 was considered statistically significant for primary analyses; exploratory findings were interpreted cautiously without multiplicity adjustment. Global Schoenfeld tests showed no evidence of violation for the full model (P=0.61) or the seven-point score (P=0.64).

Sample size was determined by the available consecutive cohort without an a priori calculation. The primary model included 75 events across four coefficients (18.75 events per coefficient). Analyses used Stata 19 and R 4.6.1, including survival, survminer and ggplot2.

## RESULTS

### Study Population and Baseline Characteristics

Among the 341 score-evaluable development recipients, 75 (22.0%) died within one year, compared with 5 of 16 (31.3%) non-evaluable recipients (HR 1.46, 95% CI 0.59–3.60; P=0.415). The small non-evaluable group limits the precision of this comparison. Baseline demographic, clinical, haemodynamic and donor characteristics, together with univariable associations with one-year mortality, are presented in Table 1.

**Table 1.**
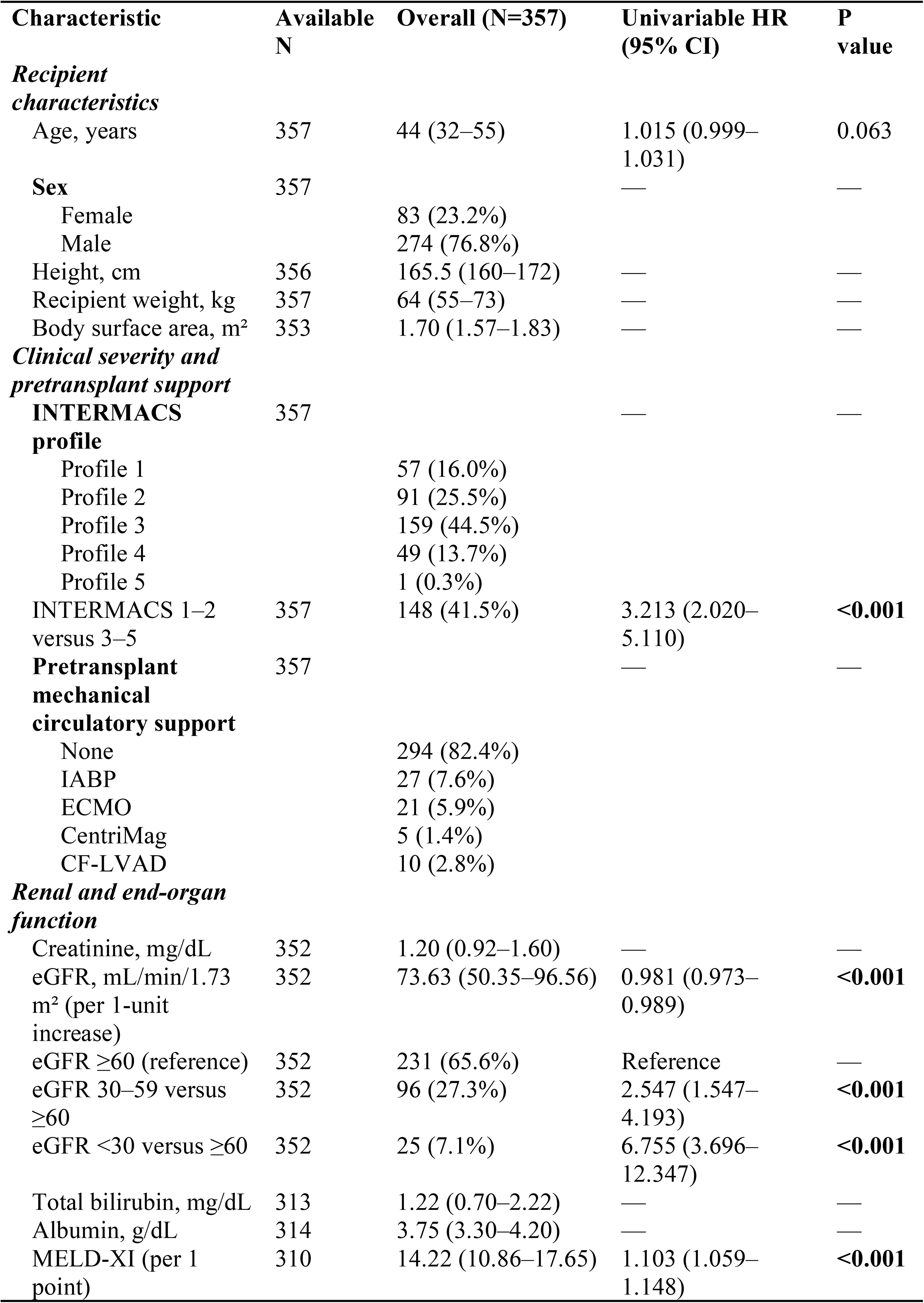
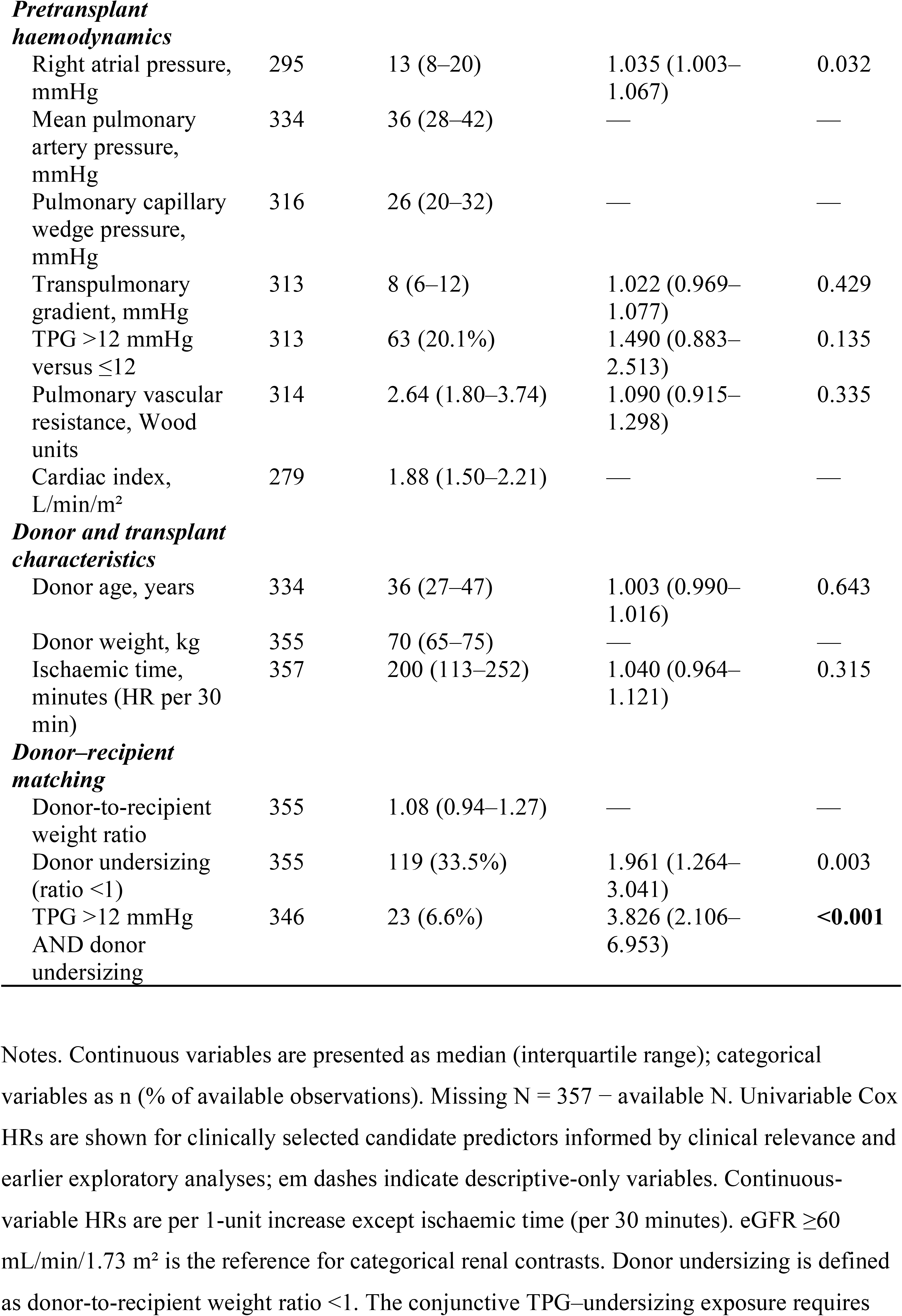
Baseline characteristics and univariable Cox associations with one-year.

| Characteristic | Available N | Overall (N=357) | Univariable HR (95% CI) | P value |
|---|---|---|---|---|
| ***Recipient characteristics*** |  |  |  |  |
| Age, years | 357 | 44 (32–55) | 1.015 (0.999–1.031) | 0.063 |
| **Sex** | 357 |  | — | — |
| Female |  | 83 (23.2%) |  |  |
| Male |  | 274 (76.8%) |  |  |
| Height, cm | 356 | 165.5 (160–172) | — | — |
| Recipient weight, kg | 357 | 64 (55–73) | — | — |
| Body surface area, m² | 353 | 1.70 (1.57–1.83) | — | — |
| ***Clinical severity and pretransplant support*** |  |  |  |  |
| **INTERMACS profile** | 357 |  | — | — |
| Profile 1 |  | 57 (16.0%) |  |  |
| Profile 2 |  | 91 (25.5%) |  |  |
| Profile 3 |  | 159 (44.5%) |  |  |
| Profile 4 |  | 49 (13.7%) |  |  |
| Profile 5 |  | 1 (0.3%) |  |  |
| INTERMACS 1–2 versus 3–5 | 357 | 148 (41.5%) | 3.213 (2.020–5.110) | **<0.001** |
| **Pretransplant mechanical circulatory support** | 357 |  | — | — |
| None |  | 294 (82.4%) |  |  |
| IABP |  | 27 (7.6%) |  |  |
| ECMO |  | 21 (5.9%) |  |  |
| CentriMag |  | 5 (1.4%) |  |  |
| CF-LVAD |  | 10 (2.8%) |  |  |
| ***Renal and end-organ function*** |  |  |  |  |
| Creatinine, mg/dL | 352 | 1.20 (0.92–1.60) | — | — |
| eGFR, mL/min/1.73 m² (per 1-unit increase) | 352 | 73.63 (50.35–96.56) | 0.981 (0.973–0.989) | **<0.001** |
| eGFR ≥60 (reference) | 352 | 231 (65.6%) | Reference | — |
| eGFR 30–59 versus ≥60 | 352 | 96 (27.3%) | 2.547 (1.547–4.193) | **<0.001** |
| eGFR <30 versus ≥60 | 352 | 25 (7.1%) | 6.755 (3.696–12.347) | **<0.001** |
| Total bilirubin, mg/dL | 313 | 1.22 (0.70–2.22) | — | — |
| Albumin, g/dL | 314 | 3.75 (3.30–4.20) | — | — |
| MELD-XI (per 1 point) | 310 | 14.22 (10.86–17.65) | 1.103 (1.059–1.148) | **<0.001** |
| ***Pretransplant haemodynamics*** |  |  |  |  |
| Right atrial pressure, mmHg | 295 | 13 (8–20) | 1.035 (1.003–1.067) | 0.032 |
| Mean pulmonary artery pressure, mmHg | 334 | 36 (28–42) | — | — |
| Pulmonary capillary wedge pressure, mmHg | 316 | 26 (20–32) | — | — |
| Transpulmonary gradient, mmHg | 313 | 8 (6–12) | 1.022 (0.969–1.077) | 0.429 |
| TPG >12 mmHg versus ≤12 | 313 | 63 (20.1%) | 1.490 (0.883–2.513) | 0.135 |
| Pulmonary vascular resistance, Wood units | 314 | 2.64 (1.80–3.74) | 1.090 (0.915–1.298) | 0.335 |
| Cardiac index, L/min/m² | 279 | 1.88 (1.50–2.21) | — | — |
| ***Donor and transplant characteristics*** |  |  |  |  |
| Donor age, years | 334 | 36 (27–47) | 1.003 (0.990–1.016) | 0.643 |
| Donor weight, kg | 355 | 70 (65–75) | — | — |
| Ischaemic time, minutes (HR per 30 min) | 357 | 200 (113–252) | 1.040 (0.964–1.121) | 0.315 |
| ***Donor–recipient matching*** |  |  |  |  |
| Donor-to-recipient weight ratio | 355 | 1.08 (0.94–1.27) | — | — |
| Donor undersizing (ratio <1) | 355 | 119 (33.5%) | 1.961 (1.264–3.041) | 0.003 |
| TPG >12 mmHg AND donor undersizing | 346 | 23 (6.6%) | 3.826 (2.106–6.953) | **<0.001** |

Notes. Continuous variables are presented as median (interquartile range); categorical variables as n (% of available observations). Missing N = 357 − available N. Univariable Cox HRs are shown for clinically selected candidate predictors informed by clinical relevance and earlier exploratory analyses; em dashes indicate descriptive-only variables. Continuous-variable HRs are per 1-unit increase except ischaemic time (per 30 minutes). eGFR ≥60 mL/min/1.73 m² is the reference for categorical renal contrasts. Donor undersizing is defined as donor-to-recipient weight ratio <1. The conjunctive TPG–undersizing exposure requires [illegible]

### Univariable Associations

INTERMACS profiles 1–2 were associated with one-year mortality (HR, 3.21; 95% CI, 2.02–5.11; P<0.001). Relative to eGFR ≥60 mL/min/1.73 m2, HRs were 2.55 (95% CI, 1.55–4.19; P<0.001) for eGFR 30–59 and 6.76 (95% CI, 3.70–12.35; P<0.001) for eGFR <30.

TPG >12 mmHg considered individually was not significantly associated with mortality (HR, 1.49; 95% CI, 0.88–2.51; P=0.135). Donor undersizing considered individually was associated with mortality (HR, 1.96; 95% CI, 1.26–3.04; P=0.003), whereas the conjunctive TPG >12 mmHg plus undersizing exposure had HR 3.83 (95% CI, 2.11–6.95; P<0.001). These candidate associations informed the final multivariable model (Table 1).

### RIGHT-HTx Model Development and Risk Stratification

The final multivariable model incorporated recipient clinical acuity, renal dysfunction, and donor– recipient haemodynamic–size compatibility (Table 2). Adjusted HRs were 2.66 (95% CI, 1.64–4.33; P<0.001) for INTERMACS 1–2; 2.41 (95% CI, 1.46–3.99; P<0.001) for eGFR 30–59; 4.09 (95% CI, 2.10–7.95; P<0.001) for eGFR <30; and 2.87 (95% CI, 1.53–5.40; P=0.001) for the conjunctive TPG–undersizing exposure.

**Table 2.**
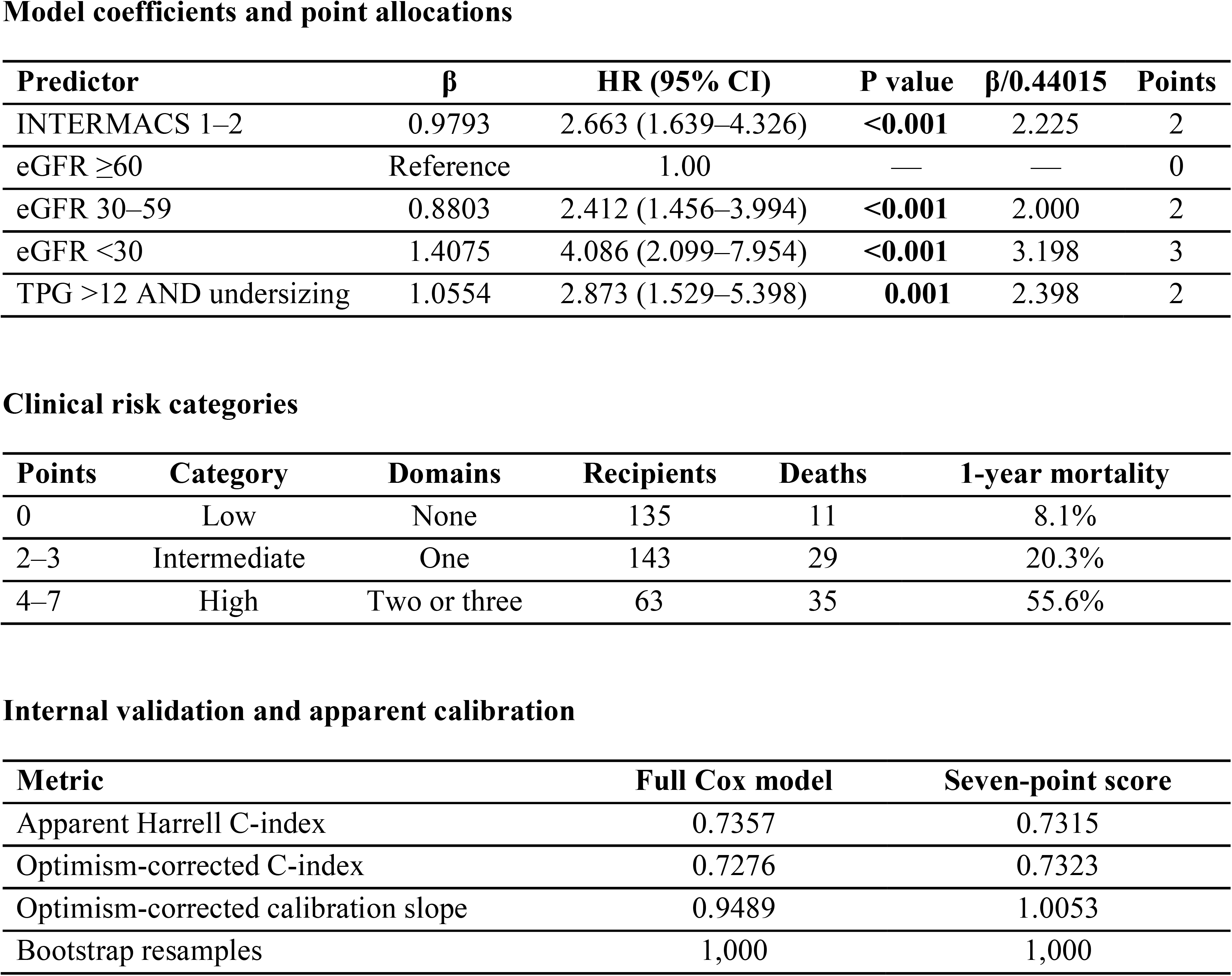
Final multivariable Cox model and derivation of the seven-point RIGHT-HTx score.

| **Model coefficients and point allocations** |  |  |  |  |  |
|---|---|---|---|---|---|
| **Predictor** | **β** | **HR (95% CI)** | **P value** | **β/0.44015** | **Points** |
| INTERMACS 1–2 | 0.9793 | 2.663 (1.639–4.326) | **<0.001** | 2.225 | 2 |
| eGFR ≥60 | Reference | 1.00 | — | — | 0 |
| eGFR 30–59 | 0.8803 | 2.412 (1.456–3.994) | **<0.001** | 2.000 | 2 |
| eGFR <30 | 1.4075 | 4.086 (2.099–7.954) | **<0.001** | 3.198 | 3 |
| TPG >12 AND undersizing | 1.0554 | 2.873 (1.529–5.398) | **0.001** | 2.398 | 2 |
| **Clinical risk categories** |  |  |  |  |  |
| **Points** | **Category** | **Domains** | **Recipients** | **Deaths** | **1-year mortality** |
| 0 | Low | None | 135 | 11 | 8.1% |
| 2–3 | Intermediate | One | 143 | 29 | 20.3% |
| 4–7 | High | Two or three | 63 | 35 | 55.6% |
| **Internal validation and apparent calibration** |  |  |  |  |  |
| **Metric** | **Full Cox model** | **Seven-point score** |  |  |  |
| Apparent Harrell C-index | 0.7357 | 0.7315 |  |  |  |
| Optimism-corrected C-index | 0.7276 | 0.7323 |  |  |  |
| Optimism-corrected calibration slope | 0.9489 | 1.0053 |  |  |  |
| Bootstrap resamples | 1,000 | 1,000 |  |  |  |

The seven-point score allocated 2 points for INTERMACS 1–2, 2 for eGFR 30–59, 3 for eGFR <30, and 2 for combined TPG >12 mmHg with undersizing. Low risk (0 points), intermediate risk (2–3), and high risk (4–7) represented zero, one, and two or more vulnerability domains.

Among 341 development recipients, 135 were low risk, 143 intermediate risk, and 63 high risk. Crude one-year mortality was 11/135 (8.1%), 29/143 (20.3%), and 35/63 (55.6%), respectively (Figure 1). Kaplan–Meier survival at one year was 91.8%, 79.7%, and 44.4%, respectively (log-rank P<0.0001; Figure 2A).

**Figure 1.**
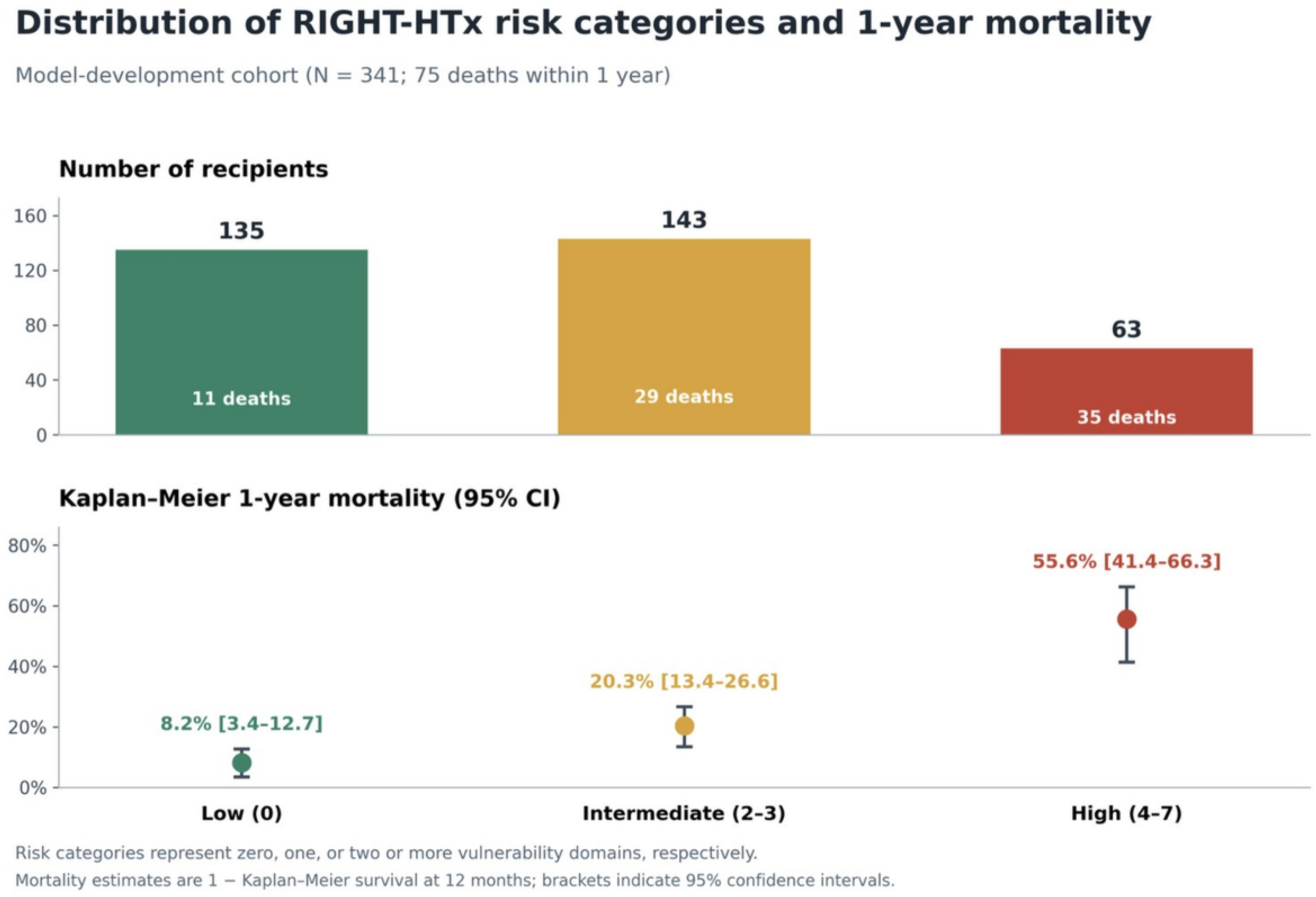
Distribution of RIGHT-HTx risk categories and one-year mortality in the model-development cohort. The upper panel shows the distribution of 341 score-evaluable heart transplant recipients across low-risk (score 0; n=135), intermediate-risk (score 2-3; n=143) and high-risk (score 4-7; n=63) categories, with the number of deaths within one year indicated. The lower panel presents Kaplan-Meier estimates of one-year mortality with 95% confidence intervals. Estimated mortality increased from 8.2% in the low-risk group to 20.3% in the intermediate-risk group and 55.6% in the high-risk group. Risk categories correspond to zero, one and two or more affected vulnerability domains, respectively.

**Figure 2.**
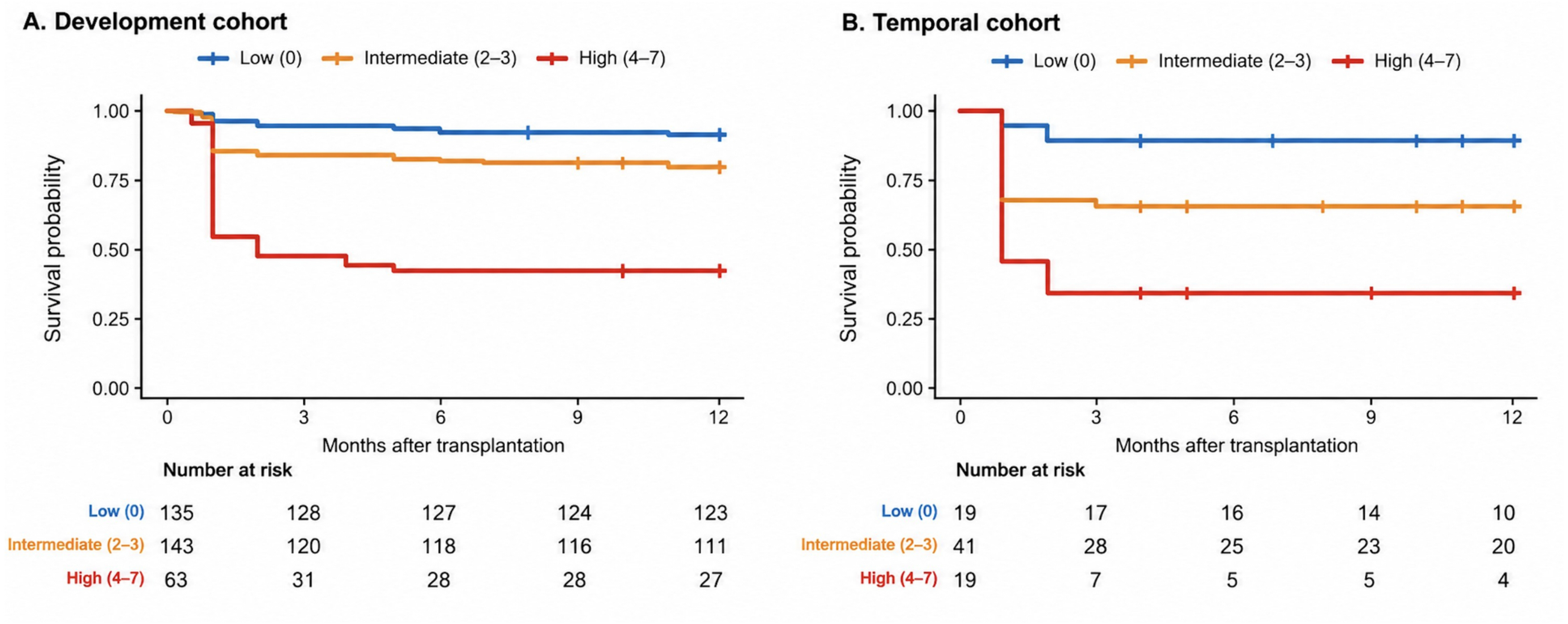
Kaplan-Meier survival according to RIGHT-HTx risk categories in the development and temporal cohorts. **(A)** Kaplan-Meier survival curves stratified by RIGHT-HTx risk category in the development cohort (n=341). (B) Corresponding survival curves in the non-overlapping, same-centre temporal cohort (n=79), using the unchanged seven-point score. Risk categories were defined as low (score 0), intermediate (scores 2-3) and high (scores 4-7), representing zero, one and two or more affected vulnerability domains, respectively. Survival differed significantly across categories in both the development (log-rank P<0.001) and temporal (log-rank P=0.003) cohorts. Tick marks indicate censored observations. Numbers at risk are displayed below each panel.

### Incremental Prognostic Value

The full three-domain model improved discrimination beyond renal dysfunction alone (apparent C-index 0.7357 versus 0.6599). In a paired bootstrap comparison using 1,000 resamples, the optimism-corrected C-index increased from 0.6594 for the renal-only model to 0.7261 for the full model (ΔC=0.0667). Adding the haemodynamic–donor domain to renal dysfunction and INTERMACS improved model fit (likelihood-ratio χ2=8.795, 1 df; P=0.003; Supplementary Table S1).

In a sensitivity analysis of 242 recipients with both measures available (58 one-year deaths), MELD-XI was associated with mortality when considered alone (HR per point 1.10; P<0.001), but not after adjustment for eGFR categories (HR 1.03; P=0.486). Adding MELD-XI to eGFR did not improve model fit (LR χ2=0.48; P=0.491), whereas adding eGFR to MELD-XI did (LR χ2=8.17; P=0.017).

### Internal Validation and Calibration

The full multivariable Cox model had an apparent Harrell C-index of 0.7357 and an optimism-corrected C-index of 0.7276 after 1,000 bootstrap resamples, with an optimism-corrected calibration slope of 0.9489. The simplified seven-point RIGHT-HTx score had an apparent C-index of 0.7315 and an optimism-corrected C-index of 0.7323, with an optimism-corrected calibration slope of 1.0053.

The slightly higher optimism-corrected estimates for the simplified score reflect differences in estimated optimism, not demonstrated superiority. Mean predicted one-year mortality in the development cohort was 21.92%, compared with observed Kaplan–Meier mortality of 22.01% (Table 2; Figure 3A). Bootstrap validation held predictors, thresholds, integer score weights and risk-category boundaries fixed and did not repeat the full model-development process.

**Figure 3.**
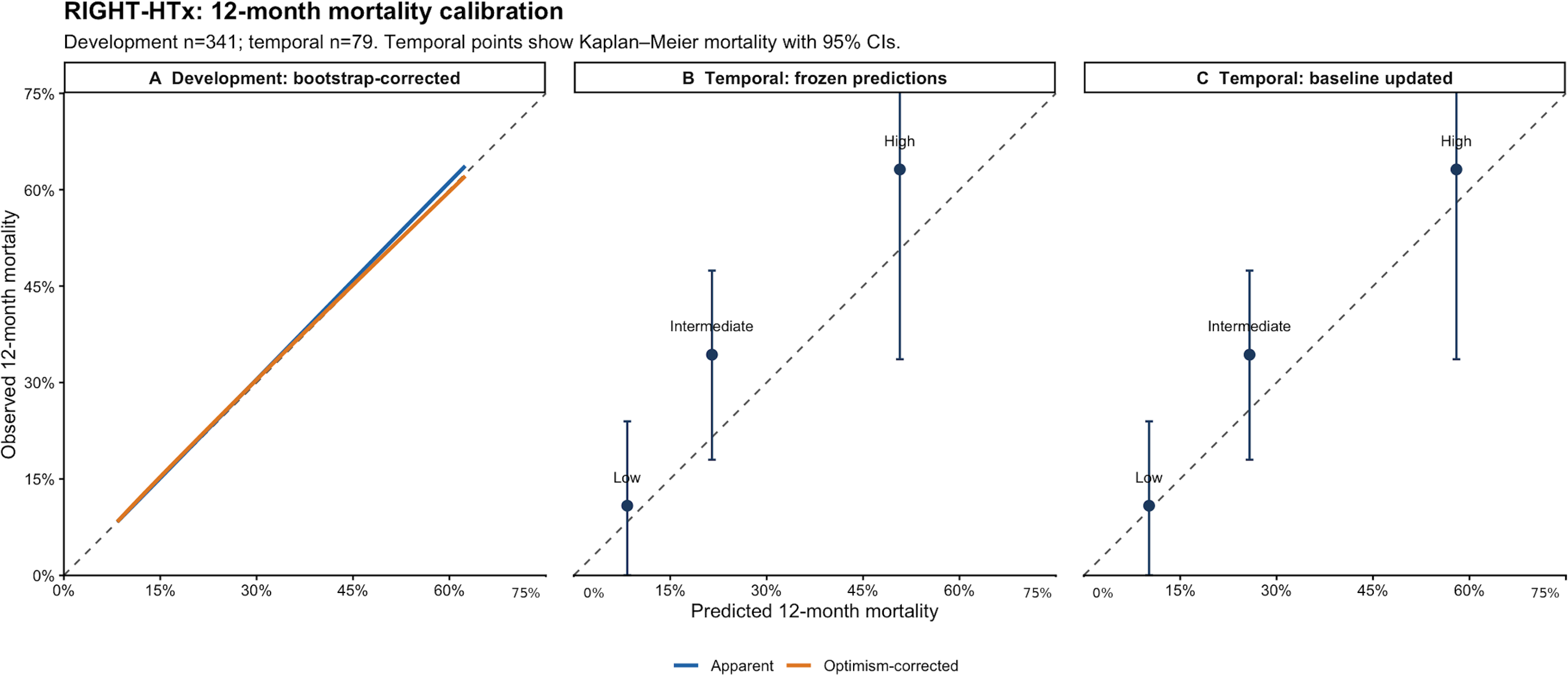
Calibration of predicted 12-month mortality in the development and temporal RIGHT-HTx cohorts. **(A)** Apparent and optimism-corrected calibration curves in the development cohort (n=341), generated using Hare smoothing and 1,000 bootstrap resamples. (B) Observed Kaplan-Meier 12-month mortality by risk category compared with predictions from the unchanged development model in the temporal cohort (n=79). (C) Observed mortality compared with predictions following exploratory updating of the baseline cumulative hazard while retaining the original score coefficient. Vertical error bars represent 95% confidence intervals for observed Kaplan-Meier mortality. The dashed diagonal indicates perfect calibration. Baseline updating used outcomes from the temporal cohort and therefore does not constitute independent validation.

### Haemodynamic–Donor Interaction and Physiological Analysis

Among 312 recipients with complete TPG and donor–recipient weight measurements, one-year mortality was 56.5% (13/23) when elevated TPG and donor undersizing coexisted, compared with 20.8% (60/289) among those with neither or only one exposure. Relative to recipients with neither exposure, the combined-exposure group had substantially higher mortality (HR 3.85, 95% CI 2.02– 7.35; P<0.001; Supplementary Table S2).

In an exploratory analysis adjusted for INTERMACS status and renal dysfunction, the multiplicative interaction between elevated TPG and donor undersizing remained statistically significant (HR 3.24, 95% CI 1.03–10.22; P=0.045; Supplementary Table S3). This finding supports the physiological rationale for the conjunctive haemodynamic–donor domain used in RIGHT-HTx, although the interaction and conjunctive models are not statistically equivalent. The interaction estimate was imprecise, with only 23 recipients having both exposures.

An analogous analysis using PVR >3 Wood units demonstrated a similar pattern, with an adjusted interaction HR of 2.66 (95% CI 1.01–7.02; P=0.049; Supplementary Table S4). Because PVR incorporates TPG and cardiac output, this represents physiological consistency rather than independent confirmation.

### Donor–Recipient Weight Matching

Among 355 development-era recipients with complete donor and recipient body-weight measurements, donor weight increased by only 1.53 kg (95% CI 0.91–2.14 kg) for every 10-kg increase in recipient weight. The fitted slope was significantly below that expected with proportional donor–recipient weight matching (P<0.001; R2=0.063). Donor undersizing increased progressively across recipient-weight categories, from 2.1% (1/48) among recipients weighing <50 kg to 98.0% (48/49) among those weighing ≥80 kg. Median donor weight increased from 65.0 to 74.8 kg across these strata, compared with an increase in median recipient weight from 45.0 to 86.0 kg. These findings demonstrate a marked disparity between recipient and donor body weights, particularly among larger recipients (Figure 4; Supplementary Table S5).

**Figure 4.**
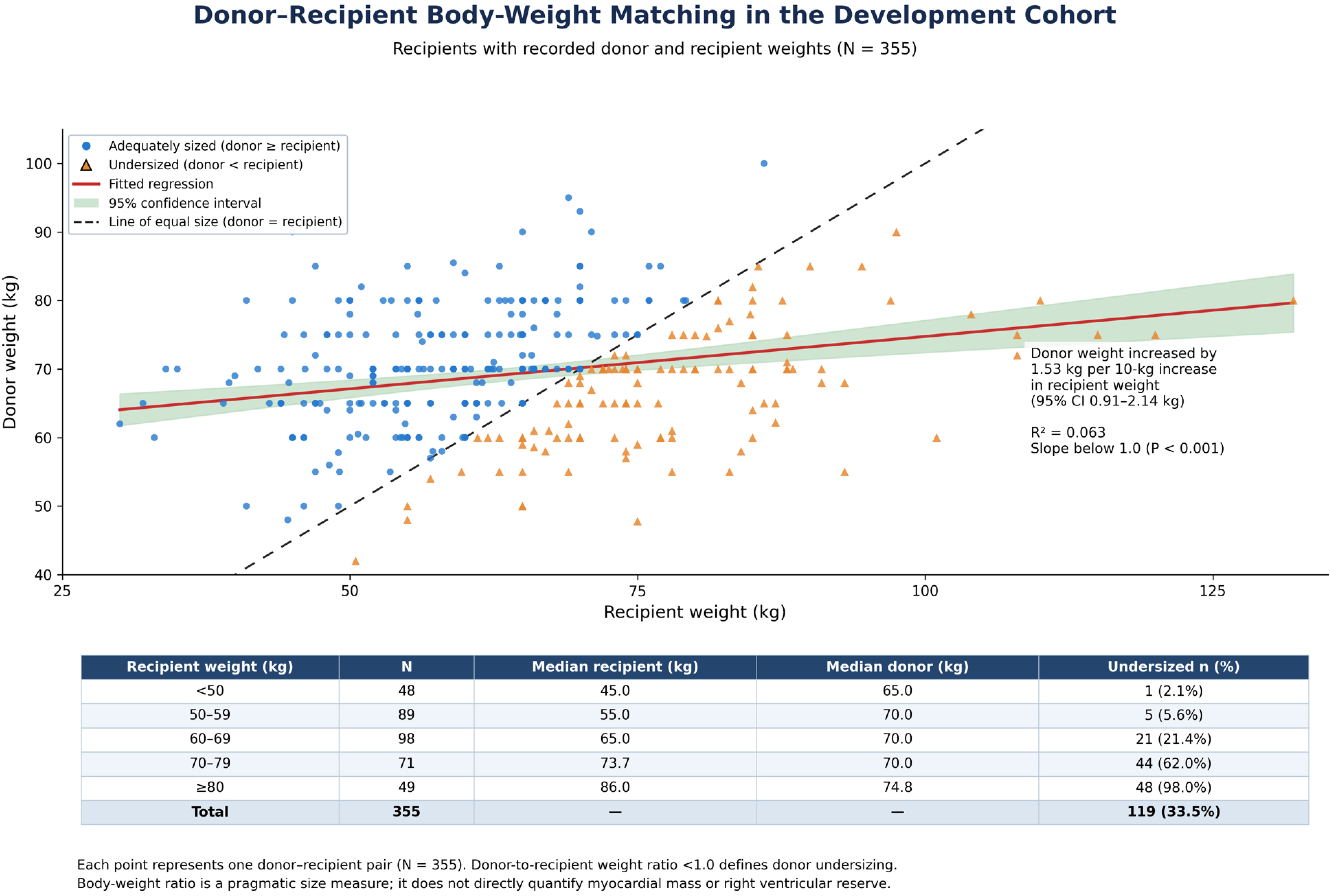
Donor-recipient body-weight matching in the RIGHT-HTx development cohort. Scatterplot of donor and recipient body weights among 355 heart transplant recipients. Blue circles represent adequately sized donor-recipient pairs (donor weight ≥ recipient weight), and orange triangles indicate donor undersizing (donor-to-recipient weight ratio <1.0). The red line represents the fitted linear regression, with the shaded region indicating its 95% confidence interval. The dashed diagonal represents equal donor and recipient weights. Donor weight increased by 1.53 kg (95% CI, 0.91-2.14 kg) for every 10-kg increase in recipient weight (R^2^=0.063). The accompanying table demonstrates progressively increasing donor undersizing across recipient­weight categories, reaching 98.0% (48/49) among recipients weighing ≥80 kg. The observed relationship describes transplanted donor-recipient pairs and does not establish donor-pool availability. Body-weight ratio is a pragmatic measure of size matching and does not directly quantify donor cardiac mass or right ventricular reserve.

### Temporal Evaluation of RIGHT-HTx

All 79 recipients in the subsequent same-centre temporal cohort were score-evaluable. Compared with the development cohort, the temporal cohort had a significantly higher prevalence of at least one represented pretransplant vulnerability (75.9% versus 60.4%; P=0.010) and higher one-year mortality (35.4% versus 22.0%). Mortality increased across RIGHT-HTx risk categories, from 10.5% in low-risk to 34.1% in intermediate-risk and 63.2% in high-risk recipients (Table 3; Figure 2B).

**Table 3.** RIGHT-HTx risk-category distribution and crude one-year mortality by cohort.

| RIGHT-HTx risk category | Development cohort, n (%) | Temporal cohort, n (%) | Development mortality | Temporal mortality |
|---|---|---|---|---|
| Low (0) | 135 (39.6%) | 19 (24.1%) | 11/135 (8.1%) | 2/19 (10.5%) |
| Intermediate (2–3) | 143 (41.9%) | 41 (51.9%) | 29/143 (20.3%) | 14/41 (34.1%) |
| High (4–7) | 63 (18.5%) | 19 (24.1%) | 35/63 (55.6%) | 12/19 (63.2%) |
| Total | 341 (100%) | 79 (100%) | 75/341 (22.0%) | 28/79 (35.4%) |

Note. Percentages in cohort columns are proportions of the respective score-evaluable cohort; mortality values are crude death proportions, not Kaplan–Meier estimates. The two cohorts are non-overlapping. A greater prevalence of represented vulnerability does not, by itself, explain all temporal differences in mortality.

The continuous seven-point RIGHT-HTx score showed discrimination comparable in magnitude to its development performance (Harrell C-index 0.738, 95% CI 0.643–0.833). The temporal calibration slope was 0.929 (95% CI 0.452–1.406).

Observed Kaplan–Meier one-year mortality exceeded predictions derived from the development model (35.44% versus 25.35%). Exploratory baseline-hazard recalibration increased mean predicted mortality to 29.56%, although underprediction remained (Figures 3B–C). Detailed calibration estimates are provided in Supplementary Table S6.

### Cumulative Vulnerability in the Pooled Cohort

Among 420 score-evaluable recipients (103 one-year deaths), mortality increased progressively with the number of affected vulnerability domains, from 8.4% with none to 23.4% with one, 54.1% with two and 87.5% (7/8) with all three. Recipients with two or more affected domains constituted only 19.5% of the cohort but accounted for 45.6% of one-year deaths (Figure 5). These pooled analyses were descriptive and did not constitute independent validation.

**Figure 5.**
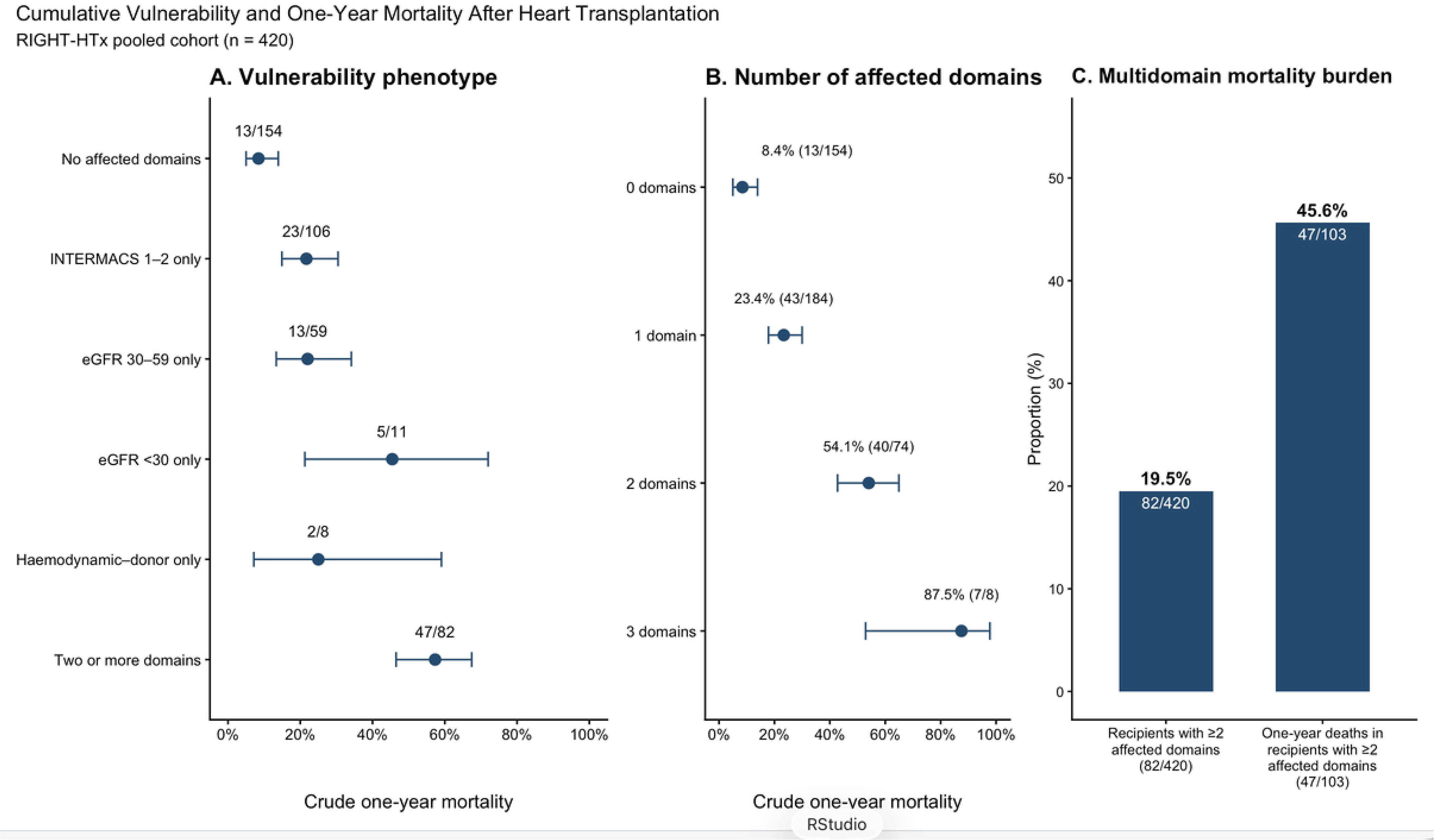
**Cumulative vulnerability, phenotype-specific mortality and distribution of one-year deaths in the pooled RIGHT-HTx cohort.** (A) Crude one-year mortality according to the type and severity of affected vulnerability domains among 420 score-evaluable recipients. Individual phenotypes are shown separately for clinical acuity (INTERMACS 1 −2), moderate renal dysfunction (eGFR 30-59 mL/min/1.73 m^2^), severe renal dysfunction (eGFR <30 mL/min/1.73 m^2^) and haemodynamic­donor mismatch (TPG >12 mmHg combined with donor undersizing). Recipients with two or more affected domains are presented as a combined category. (B) Crude one-year mortality according to the number of affected domains (zero to three), with renal dysfunction counted as one domain regardless of severity. (C) Proportion of recipients with two or more affected domains (82/420; 19.5%) compared with their contribution to all recorded one-year deaths (47/103; 45.6%). Points represent crude mortality proportions, and horizontal error bars indicate 95% Wilson binomial confidence intervals. The pooled analyses are descriptive, include development and temporal recipients, and do not constitute independent validation.

### Programme-Level and Conditional Long-Term Survival

Among 436 adult heart transplant recipients, 141 deaths occurred during a median follow-up of 4.7 years (IQR, 2.0–8.0 years), of which 108 (76.6%) occurred within the first post-transplant year.

Overall Kaplan–Meier survival was 75.1% at one year, 68.2% at five years and 57.7% at ten years (Supplementary Figure S1A)

Among 341 score-evaluable development recipients, 261 survived and remained under observation beyond the one-year landmark, with 31 subsequent deaths. Conditional survival was 91.0% at five years and 76.7% at ten years after transplantation (Supplement Figure S1B).

Among these one-year survivors, subsequent survival did not differ significantly across the original RIGHT-HTx risk categories. Five-year survival was 92.3%, 89.0% and 90.7% in low-, intermediate- and high-risk recipients, respectively (log-rank P=0.60; C-index 0.515; Supplement Figure S1C).

These findings suggest that the prognostic separation associated with RIGHT-HTx is concentrated predominantly within the first post-transplant year, although the limited number of late deaths and selection inherent in landmark analysis preclude conclusions of equivalent long-term risk.

## DISCUSSION

Risk models derived from high-income registries may not transport readily to resource-constrained programmes, where recipient acuity, donor characteristics, and access to advanced bridging differ. RIGHT-HTx organised transplant risk into three domains: clinical acuity, renal reserve, and donor– recipient haemodynamic compatibility. One-year survival in the development cohort ranged from 91.8% in low-risk to 44.4% in high-risk recipients. Risk separation persisted in subsequent recipients despite greater pretransplant vulnerability and higher mortality (C-statistic 0.738). The principal contribution of RIGHT-HTx may lie more in demonstrating the convergence of vulnerabilities than in the numerical risk score alone, although sparse phenotypes cannot establish synergy.

Clinical acuity,^20^ renal dysfunction,^21,22^ pulmonary vascular load^23^ and donor–recipient size mismatch^24^ are established determinants of heart-transplant outcomes.

Risk factors seldom operate in isolation, and cumulative vulnerability, including comorbidity burden,^25^ frailty and obesity,^26^ and cardiovascular–kidney–metabolic risk,^27^ has been associated with post-transplant mortality. Previous models have incorporated combinations of donor and recipient risk factors,^28,29^ with analogous approaches in lung^30^ and liver^31^ transplantation. However, the cumulative and potentially interrelated effects of these physiological vulnerabilities remain incompletely characterised. We are not aware of an existing framework that integrates these three domains and evaluates their combined prognostic contribution in a subsequent temporal cohort.

The donor–recipient haemodynamic domain illustrates the relational nature of RIGHT-HTx: elevated TPG was most informative when combined with donor undersizing, consistent with previous evidence that donor and recipient characteristics may modify each other’s effects.^28,32^ One-year mortality was 56.5% when both exposures coexisted, compared with 20.8% among recipients with neither or only one exposure. A similar pattern was observed with PVR, supporting the physiological rationale for considering pulmonary vascular load in relation to donor size rather than in isolation.^33,34^

RIGHT-HTx showed similar discrimination in the subsequent temporal cohort (C-index 0.738) despite greater pretransplant vulnerability. Observed mortality exceeded predictions from the development model, highlighting the distinction between discrimination and absolute-risk calibration.^35,36^ Exploratory baseline-hazard recalibration only partially reduced this discrepancy, underscoring the need for independent validation and potential recalibration across different populations.^37^ During the later period, the programme also experienced disruptions in air transport, increased retrieval costs and difficulties with timely donor-organ procurement, potentially affecting waiting times and clinical status at transplantation. These circumstances illustrate the challenges of transplantation in a resource-constrained setting, but their contribution to the higher mortality could not be determined from the available data, as the relevant variables — waiting-list duration, retrieval disruptions and their timing — were not recorded.

Two deaths among low-risk recipients in the subsequent cohort illustrate vulnerabilities not adequately captured by RIGHT-HTx. One involved complex congenital heart disease after Fontan palliation and multiple previous operations; the other involved PVR 5.3 Wood units and TPG 14 mmHg without meeting the predefined donor-undersizing criterion (donor 70 kg; recipient 67 kg). These cases highlight an inherent limitation of parsimonious, threshold-based risk scores: a score of zero does not imply absence of clinical risk.

Donor-size availability emerged as an important systems-level constraint in our transplant programme. Whereas previous matching studies have examined acceptable mismatch and organ utilisation,^24,38,39,40^ our data demonstrate relatively constant donor weight across increasing recipient size, with undersizing increasing from 2.1% among recipients weighing <50 kg to 98.0% among those weighing ≥80 kg. Median recipient weight increased from 45.0 to 86.0 kg across these strata, whereas median donor weight increased only from 65.0 to 74.8 kg. These observations demonstrate the practical constraints of donor–recipient matching among transplanted recipients, although they do not establish the availability of appropriately sized hearts in the underlying donor pool. Size mismatch may represent a continuous gradient rather than a discrete threshold.40 Predicted heart mass (PHM) may provide more precise size matching than body weight,^41,42^ although ideal body-weight ratio has been reported non-inferior to PHM.^43^ PHM could not be reliably calculated in most recipients because of incomplete donor anthropometric data. Importantly, neither metric directly measures donor right ventricular reserve relative to recipient pulmonary vascular load. The RIGHT-HTx weight-ratio threshold should therefore be regarded as a pragmatic operational definition rather than a physiological boundary.

The healthcare-system context is important when interpreting transplant outcomes. The two principal surgical options for end-stage heart failure—heart transplantation and durable LVAD support—depend on resources constrained by different factors: organ donation rates and access to reimbursement, respectively. Registry-derived risk scores, such as IMPACT, incorporate treatment-dependent variables, including mechanical ventilation and circulatory support, whose prognostic significance may differ across healthcare systems.^44,45^ In resource-constrained programmes, the absence of advanced support may reflect limited access or affordability rather than lesser clinical need. Such variables may therefore capture both disease severity and healthcare resources, potentially limiting the transportability of existing scores.46 RIGHT-HTx instead used transplant-time INTERMACS profile to characterise clinical acuity less directly dependent on the support modality received. Direct benchmarking against established scores was not undertaken because the available data did not permit reliable reconstruction of all comparator predictors and some scores target different endpoints.

The cumulative-vulnerability framework also has limitations. Severity within a domain matters, not merely the number of affected domains. Recipients with multiple vulnerabilities constituted only 19.5% of the cohort but accounted for 45.6% of one-year deaths. However, the adverse prognosis associated with severe renal dysfunction illustrates that the severity of individual vulnerabilities may be as important as their accumulation.

RIGHT-HTx was developed to identify potentially modifiable peri-transplant vulnerability rather than determine transplant eligibility or organ allocation. This emphasis on early vulnerability is consistent with its prognostic separation being concentrated predominantly within the first post-transplant year, with no significant risk-category separation among one-year survivors (log-rank P=0.60).

Its three domains suggest different opportunities for intervention. Earlier referral, before deterioration in renal function and clinical acuity, may help preserve physiological reserve.^47,48^ Timely mechanical circulatory support may prevent progression to critical illness,^49^ while recognition of elevated pulmonary vascular load may inform donor–recipient size matching. Reversible pulmonary vascular load may also be modified through durable mechanical circulatory support,^50^ yet access to affordable devices remains limited in this setting, where treatment costs are largely borne by recipients.^8,9,10^ These are potential strategies rather than interventions evaluated in this study, and prospective evaluation is needed to determine whether score-informed management improves outcomes. Favourable outcomes among low-risk recipients highlight the potential importance of reaching transplantation before multiple vulnerabilities accumulate, while residual mortality even at score zero emphasises that low scores do not exclude clinical risk.

### LIMITATIONS

This retrospective study reflects the consecutive experience of a single transplant programme. Its findings describe early post-transplant risk within that setting and require evaluation in other populations.

Score calculation was possible in 341 of 357 development recipients (95.5%). Among the 16 non-evaluable recipients, five died within one year (31.3%), compared with 75 of 341 (22.0%) evaluable recipients; their exclusion may therefore have introduced selection bias. Eleven recipients had indeterminate haemodynamic–donor classification: nine had donor undersizing with missing TPG, and two lacked donor-weight measurements. Multiple imputation was not undertaken. The primary model contained 75 deaths across four coefficients, and predictor selection, thresholds and integer weights were informed by development data. Although risk-category boundaries followed the number of affected domains, bootstrap validation was conditional on these modelling decisions and may underestimate full-development optimism.

The adjusted TPG interaction analysis included only 23 recipients with both exposures, producing an imprecise exploratory estimate. PVR provided physiological consistency rather than independent replication. Haemodynamic measurements are influenced by loading conditions and timing, while body weight is an imperfect surrogate for donor cardiac mass and right ventricular reserve. The observed donor–recipient matching pattern describes transplanted recipients and cannot establish donor-pool availability.

The 79-recipient temporal cohort provided limited precision for calibration and risk-category comparisons. Seventeen recipients had less than 12 months of observation, although none was lost to follow-up. The temporal evaluation was conducted at the same centre and does not constitute geographical external validation. Baseline-hazard recalibration used temporal outcomes and did not eliminate the discrepancy between predicted and observed mortality. Small pooled phenotypes further limit precision.

Only 31 late deaths occurred among development recipients surviving beyond one year, limiting interpretation of conditional survival comparisons. Important potential predictors—including detailed graft quality, cause-specific mortality, waiting-list deterioration, donor availability, treatment access and financial constraints—were incompletely captured. A score of zero indicates the absence of vulnerabilities represented in RIGHT-HTx, not the absence of clinical risk; complex congenital anatomy, allosensitisation and other unmeasured factors may remain important. Independent multicentre validation and prospective evaluation are required before clinical implementation.

## CONCLUSIONS

RIGHT-HTx integrates recipient clinical acuity, renal dysfunction and donor–recipient haemodynamic–size compatibility into a parsimonious seven-point score for one-year mortality after adult heart transplantation. Its categories correspond to zero, one or multiple score-captured vulnerability domains. Discrimination was comparable in magnitude in a later, more vulnerable same-centre cohort, although absolute-risk calibration differed. The framework may help recognise potentially modifiable vulnerability but should not be used to exclude patients from transplantation or determine organ allocation without independent evaluation.

## Data Availability

The individual patient-level data underlying this study are not publicly available because of patient confidentiality, institutional data governance requirements and the potential for re-identification. Requests for access may be considered by the corresponding author, subject to institutional approval and applicable ethical and legal requirements.

---

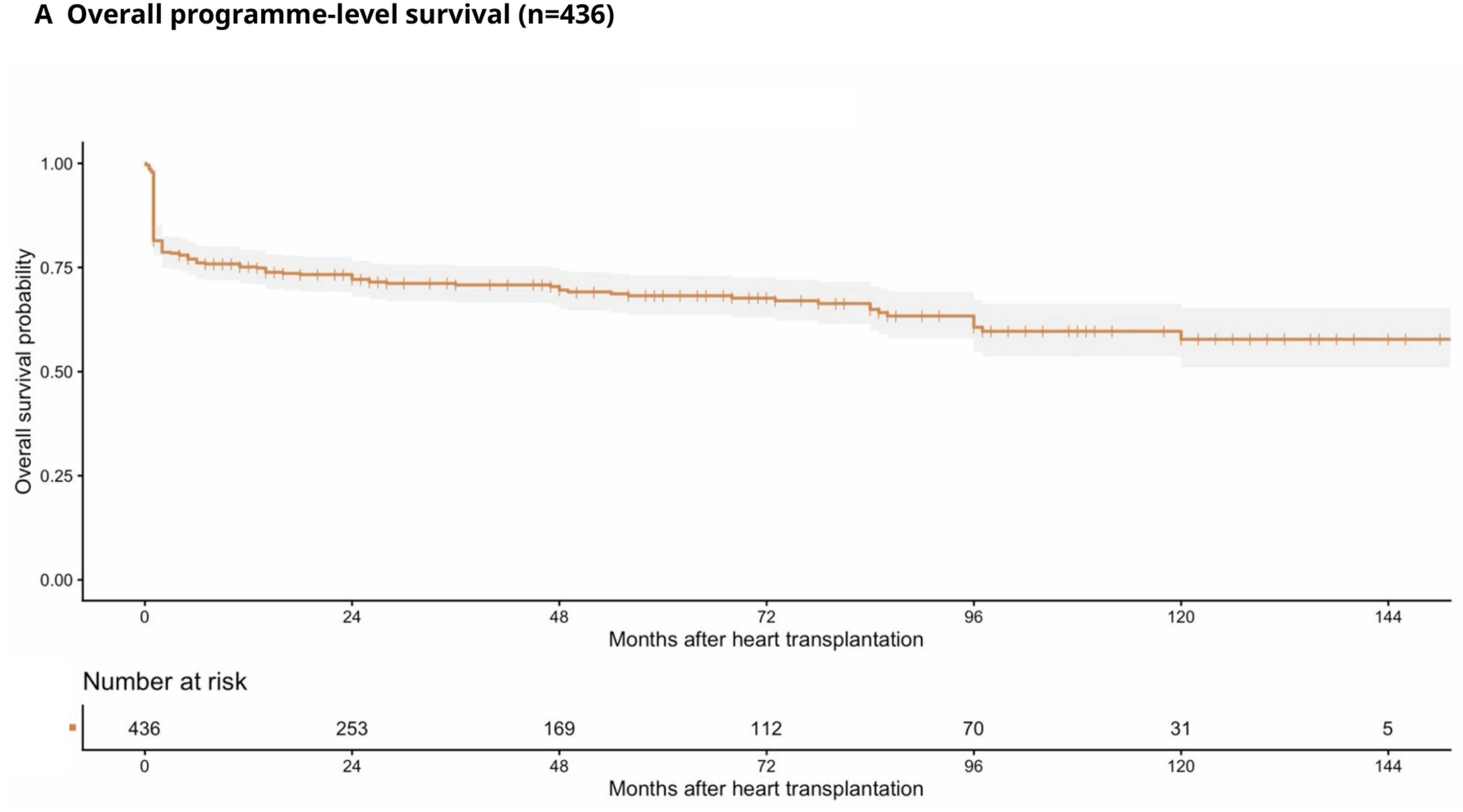

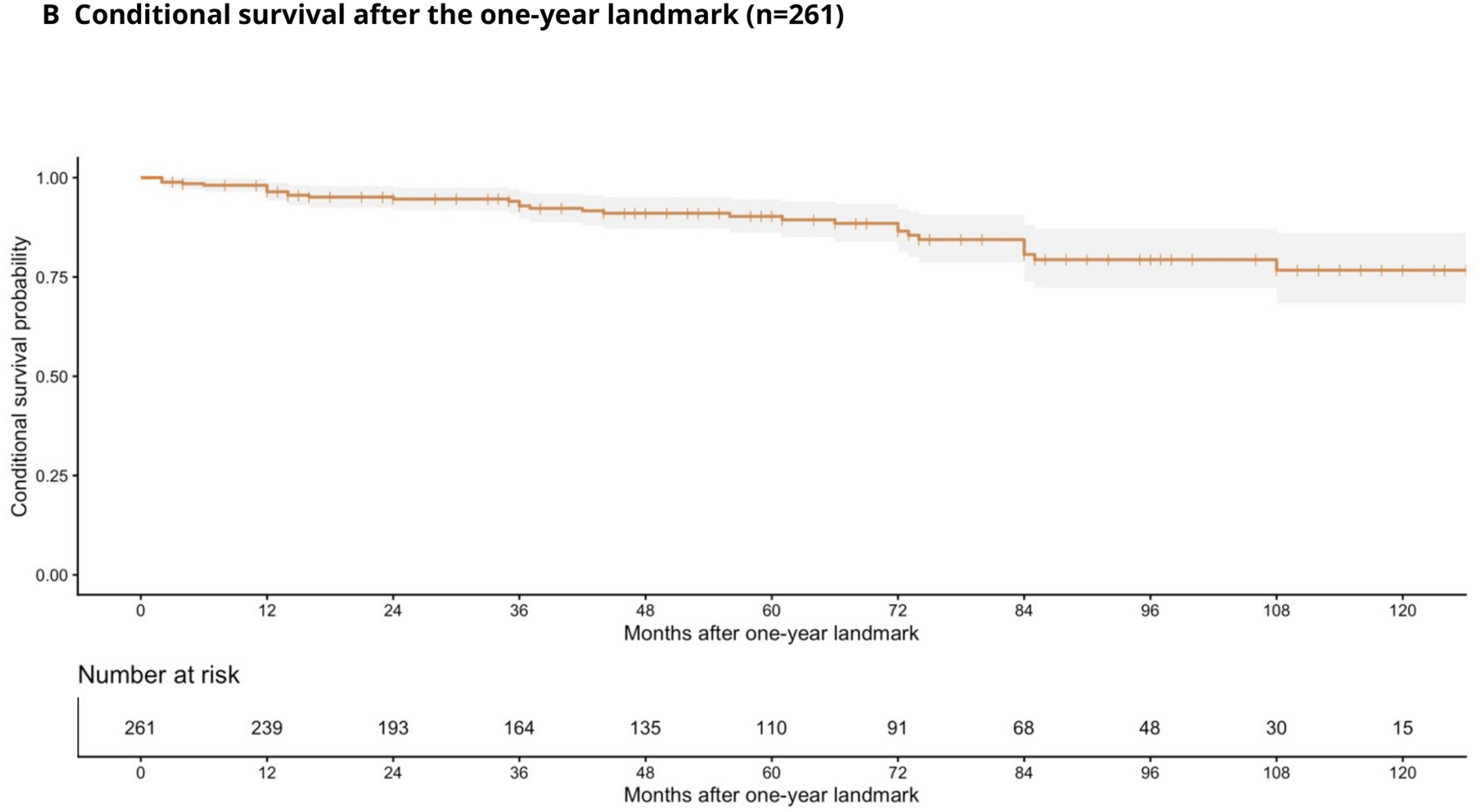

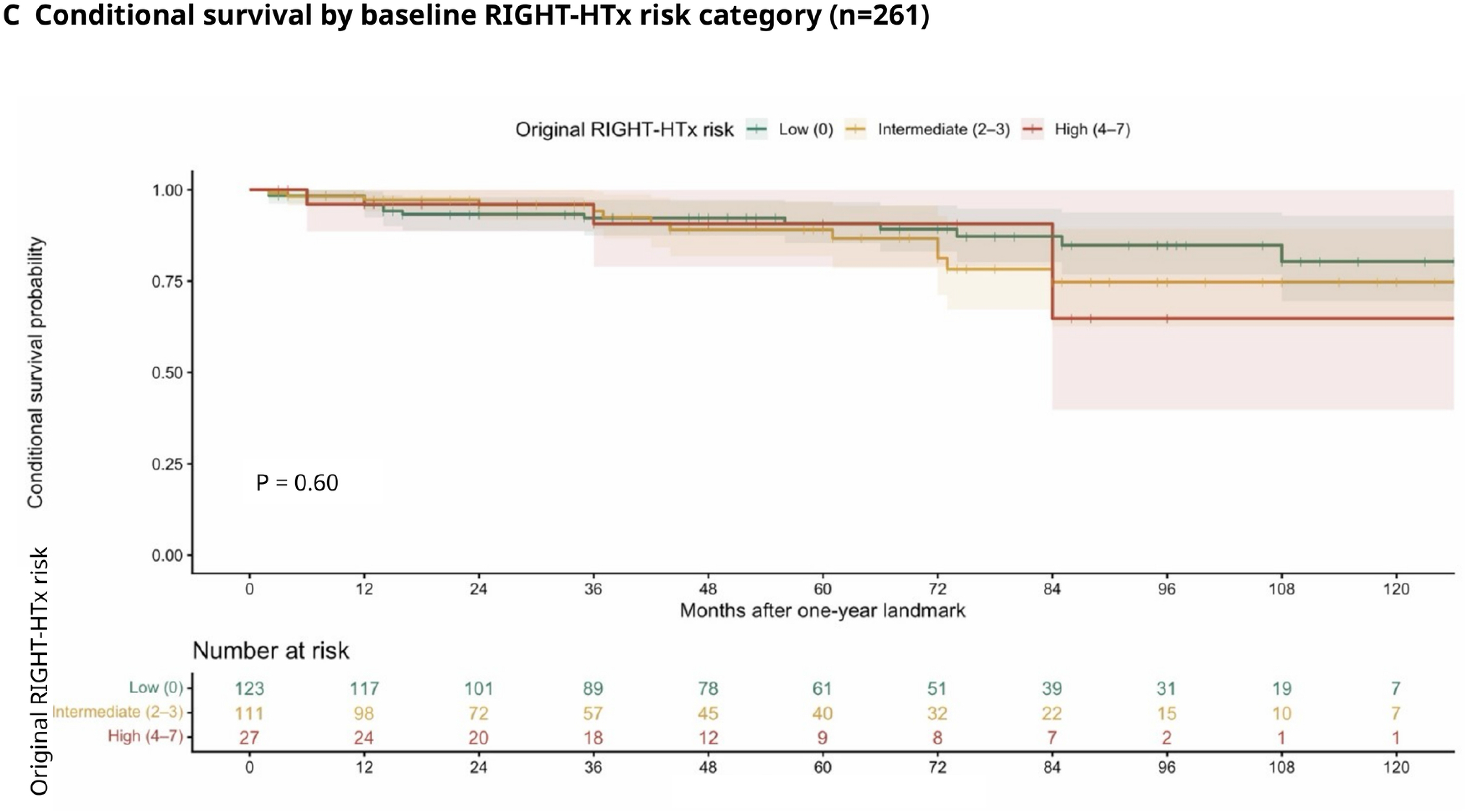

## References

1. Severino P, Mather PJ, Pucci M, et al. Advanced heart failure and end-stage heart failure: does a difference exist? Diagnostics (Basel). 2019;9(4):170.

2. El Rafei A, Cogswell R, Atik FA, Zuckermann A, Allen LA. Review of the global activity of heart transplant. Circ Heart Fail. 2025;18(7):e012272.

3. Ortiz-Bautista C, Muñiz J, Almenar-Bonet L, et al. Utility of the IMPACT score for predicting heart transplant mortality: analysis on a contemporary cohort of the Spanish Heart Transplant Registry. Clin Transplant. 2022;36(9):e14774.

4. Segovia J, Cosío MDG, Barceló JM, et al. RADIAL: a novel primary graft failure risk score in heart transplantation. J Heart Lung Transplant. 2011;30(6):644–651.

5. Collins GS, de Groot JA, Dutton S, et al. External validation of multivariable prediction models: a systematic review of methodological conduct and reporting. BMC Med Res Methodol. 2014;14:40.

6. Binuya MAE, Engelhardt EG, Schats W, Schmidt MK, Steyerberg EW. Methodological guidance for the evaluation and updating of clinical prediction models: a systematic review. BMC Med Res Methodol. 2022;22(1):316.

7. Coutance G, Kransdorf E, Bonnet G, Loupy A, Kobashigawa J, Patel J. Statistical performance of 16 posttransplant risk scores in a contemporary cohort of heart transplant recipients. Am J Transplant. 2021;21(2):645–656.

8. Varshney S, Kansra P, Garg A, Raj BM, Singh SP, Khadar AA. Financial implications of organ transplant and advantages of covering it in Ayushman Bharat. J Mar Med Soc. 2025;28(1):89–93.

9. Balakrishnan K. Heart transplantation in India—are we there yet? Indian J Thorac Cardiovasc Surg. 2020;36:159–165.

10. Sivathasan C, Hayward C, Jansz P, et al. Durable mechanical circulatory support across the Asia-Pacific region. J Heart Lung Transplant. 2020;39(11):1195–1198.

11. Manek P, Madkaiker A, Gandhi H, et al. Current status, challenges, and solution for heart transplantation in public sector hospital. Indian J Transplant. 2024;18(4):352–359.

12. Mentias A, Desai MY, Vaughan-Sarrazin MS, et al. Community-level economic distress, race, and risk of adverse outcomes after heart failure hospitalization among Medicare beneficiaries. Circulation. 2022;145(2):110–121.

13. Chen Q, Malas J, Emerson D, et al. Heart transplantation in patients from socioeconomically distressed communities. J Heart Lung Transplant. 2024;43(2):324–333.

14. Axelrod DA, Guidinger MK, Finlayson S, et al. Rates of solid-organ wait-listing, transplantation, and survival among residents of rural and urban areas. JAMA. 2008;299(2):202–207.

15. Collins GS, Moons KGM, Dhiman P, et al. TRIPOD+AI statement: updated guidance for reporting clinical prediction models that use regression or machine learning methods. BMJ. 2024;385:e078378.

16. Inker LA, Eneanya ND, Coresh J, et al. New creatinine- and cystatin C–based equations to estimate GFR without race. N Engl J Med. 2021;385(19):1737–1749.

17. Heuman DM, Mihas AA, Habib A, et al. MELD-XI: a rational approach to “sickest first” liver transplantation in cirrhotic patients requiring anticoagulant therapy. Liver Transpl. 2007;13(1):30–37.

18. Friesen EL, Foroutan F, Krakovsky J, et al. Utility of the INTERMACS profile at the time of assessment for heart transplant. Clin Transplant. 2020;34(3):e13796.

19. Sullivan LM, Massaro JM, D’Agostino RB Sr. Presentation of multivariate data for clinical use: the Framingham Study risk score functions. Stat Med. 2004;23(10):1631–1660.

20. Barge-Caballero E, Segovia-Cubero J, Almenar-Bonet L, et al. Preoperative INTERMACS profiles determine postoperative outcomes in critically ill patients undergoing emergency heart transplantation: analysis of the Spanish National Heart Transplant Registry. Circ Heart Fail. 2013;6(4):763–772.

21. Kumar A, Bonnell LN, Thomas CP. Impact of changing renal function, while waiting for a heart transplant, on post-transplant mortality and development of end stage kidney disease. Transpl Int. 2021;34:1044–1051.

22. Habib PJ, Patel PC, Hodge D, et al. Pre-orthotopic heart transplant estimated glomerular filtration rate predicts post-transplant mortality and renal outcomes: an analysis of the UNOS database. J Heart Lung Transplant. 2016;35(12):1471–1479.

23. Erickson KW, Costanzo-Nordin MR, O’Sullivan EJ, et al. Influence of preoperative transpulmonary gradient on late mortality after orthotopic heart transplantation. J Heart Transplant. 1990;9(5):526–537.

24. Holzhauser L, Imamura T, Bassi N, et al. Increasing heart transplant donor pool by liberalization of size matching. J Heart Lung Transplant. 2019;38(11):1197–1205.

25. Bonnesen K, Mols RE, Løgstrup BB, Gustafsson F, Eiskjær H, Schmidt M. The ability of comorbidity indices to predict mortality after heart transplantation: a validation of the Danish Comorbidity Index for Acute Myocardial Infarction, Charlson Comorbidity Index, and Elixhauser Comorbidity Index. Transplant Direct. 2023;9(4):e1438.

26. Goubar T, Mullin AM, Rushworth LR, Macdonald PS. Frailty and obesity in heart transplant recipients: impact on post-transplant mortality. Eur Heart J. 2025;46(suppl 1):ehaf784.1357.

27. Stauffacher G, Gros BP, Antiochos P, et al. Prognostic impact of cardiovascular-kidney-metabolic burden among heart transplant patients: insights from the Swiss Transplant Cohort Study. Eur Heart J. 2025;46(suppl 1):ehaf784.1062.

28. Trivedi JR, Cheng A, Ising MS, Lenneman A, Birks EJ, Slaughter MS. Heart transplant survival based on recipient and donor risk scoring: a UNOS database analysis. ASAIO J. 2016;62(3):297–301.

29. Hong KN, Iribarne A, Worku B, et al. Who is the high-risk recipient? Predicting mortality after heart transplant using pretransplant donor and recipient risk factors. Ann Thorac Surg. 2011;92(2):520–527.

30. Ehrsam JP, Schuurmans MM, Laager M, Opitz I, İnci İ. Recipient comorbidities for prediction of primary graft dysfunction, chronic allograft dysfunction and survival after lung transplantation. Transpl Int. 2022;35:10451.

31. Volk ML, Hernandez JC, Lok AS, Marrero JA. Modified Charlson comorbidity index for predicting survival after liver transplantation. Liver Transpl. 2007;13(11):1515–1520.

32. Young JB, Naftel DC, Bourge RC, et al. Matching the heart donor and heart transplant recipient. Clues for successful expansion of the donor pool: a multivariable, multiinstitutional report. The Cardiac Transplant Research Database Group. J Heart Lung Transplant. 1994;13(3):353.

33. Chen JM, Levin HR, Michler RE, Prusmack CJ, Rose EA, Aaronson KD. Reevaluating the significance of pulmonary hypertension before cardiac transplantation: determination of optimal thresholds and quantification of the effect of reversibility on perioperative mortality. J Thorac Cardiovasc Surg. 1997;114(4):627–634.

34. Wagner T, Magnussen C, Bernhardt A, et al. Impact of diastolic pulmonary gradient and pulmonary artery pulse index on outcomes in heart transplant patients—results from the Eurotransplant database. Front Cardiovasc Med. 2022;9:1036547.

35. Van Calster B, Steyerberg EW, Wynants L, van Smeden M. There is no such thing as a validated prediction model. BMC Med. 2023;21(1):70.

36. Tanner KT, Keogh RH, Coupland CAC, Hippisley-Cox J, Diaz-Ordaz K. Dynamic updating of clinical survival prediction models in a changing environment. Diagn Progn Res. 2023;7(1):24.

37. Royston P, Altman DG. External validation of a Cox prognostic model: principles and methods. BMC Med Res Methodol. 2013;13:33.

38. Patel ND, Weiss ES, Nwakanma LU, Russell SD, Baumgartner WA, Shah AS, Conte JV. Impact of donor-to-recipient weight ratio on survival after heart transplantation: analysis of the United Network for Organ Sharing database. Circulation. 2008;118(14 suppl):S83–S88.

39. Tao R, Hess T, Kuchnia AJ, Hermsen JL, Raza F, Dhingra R. Association of size matching using predicted heart mass with mortality in heart transplant recipients with obesity or high pulmonary vascular resistance. JAMA Netw Open. 2023;6(6):e2319191.

40. Dale R, Bahatyrevich N, Leipzig M, Pines KC, Currie M. Causal nonlinear dose response analysis of donor–recipient predicted heart mass ratio. JHLT Open. 2025;11:100418.

41. Ahmad D, Brodie A, Pritting C, et al. Predicted heart mass based on ideal body weight for donor-to-recipient size matching. Clin Transplant. 2023;37(12):e15150.

42. Khush KK, Cherikh WS, Chambers DC, et al. The International Thoracic Organ Transplant Registry of the International Society for Heart and Lung Transplantation: thirty-sixth adult heart transplantation report — 2019; focus theme: donor and recipient size match. J Heart Lung Transplant. 2019;38(10):1056–1066.

43. Pritting C, Nasher N, Rahimov D, et al. Ideal body weight ratio for donor-to-recipient size matching in heart transplantation. Clin Transplant. 2025;39(11):e70369.

44. Moayedi Y, Ródenas-Alesina E, Mueller B, et al. Rethinking donor and recipient risk matching in Europe and North America: using heart transplant predictors of donor and recipient risk. Circ Heart Fail. 2023;16(5):e009994.

45. Weiss ES, Allen JG, Arnaoutakis GJ, George TJ, Russell SD, Shah AS, Conte JV. Creation of a quantitative recipient risk index for mortality prediction after cardiac transplantation (IMPACT). Ann Thorac Surg. 2011;92(3):914–921.

46. Figueredo A, Echeverría LE, Castillo VR, Murcia A, Rojas LZ, Gamboa-Delgado EM. Evaluación del riesgo de mortalidad de pacientes de trasplante cardíaco a través de la escala IMPACT en una población latinoamericana. Rev Argent Cardiol. 2016;84(5):487–492.

47. Thorvaldsen T, Lund LH. Focusing on referral rather than selection for advanced heart failure therapies. Card Fail Rev. 2019;5(1):24–26.

48. Morris AA, Khazanie P, Drazner MH, et al. Guidance for timely and appropriate referral of patients with advanced heart failure: a scientific statement from the American Heart Association. Circulation. 2021;144(15):e238–e250.

49. Barge-Caballero E, Almenar-Bonet L, Gonzalez-Vilchez F, et al. Clinical outcomes of temporary mechanical circulatory support as a direct bridge to heart transplantation: a nationwide Spanish registry. Eur J Heart Fail. 2018;20(1):178–186.

50. Salzberg SP, Lachat ML, von Harbou K, Zünd G, Turina MI. Normalization of high pulmonary vascular resistance with LVAD support in heart transplantation candidates. Eur J Cardiothorac Surg. 2005;27(2):222–225.

